# RT-LAMP has high accuracy for detecting SARS-CoV-2 in saliva and naso/oropharyngeal swabs from asymptomatic and symptomatic individuals

**DOI:** 10.1101/2021.06.28.21259398

**Authors:** Stephen P. Kidd, Daniel Burns, Bryony Armson, Andrew D. Beggs, Emma L. A. Howson, Anthony Williams, Gemma Snell, Emma L. Wise, Alice Goring, Zoe Vincent-Mistiaen, Seden Grippon, Jason Sawyer, Claire Cassar, David Cross, Thomas Lewis, Scott M. Reid, Samantha Rivers, Joe James, Paul Skinner, Ashley Banyard, Kerrie Davies, Anetta Ptasinska, Celina Whalley, Jack Ferguson, Claire Bryer, Charlie Poxon, Andrew Bosworth, Michael Kidd, Alex Richter, Jane Burton, Hannah Love, Sarah Fouch, Claire Tillyer, Amy Sowood, Helen Patrick, Nathan Moore, Michael Andreou, Nick Morant, Rebecca Houghton, Joe Parker, Joanne Slater-Jefferies, Ian Brown, Cosima Gretton, Zandra Deans, Deborah Porter, Nicholas J. Cortes, Angela Douglas, Sue L. Hill, Keith M. Godfrey, Veronica L. Fowler

## Abstract

Previous studies have described RT-LAMP methodology for the rapid detection of SARS-CoV-2 in nasopharyngeal (NP) and oropharyngeal (OP) swab and saliva samples. This study describes the validation of an improved sample preparation method for extraction free RT-LAMP and defines the clinical performance of four different RT-LAMP assay formats for detection of SARS-CoV-2 within a multisite clinical evaluation. Direct RT-LAMP was performed on 559 swabs and 86,760 saliva samples and RNA RT-LAMP on extracted RNA from 12,619 swabs and 12,521 saliva from asymptomatic and symptomatic individuals across healthcare and community settings. For Direct RT-LAMP, overall diagnostic sensitivity (DSe) of 70.35% (95% CI 63.48-76.60%) on swabs and 84.62% (79.50-88.88%) on saliva was observed, with diagnostic specificity (DSp) of 100% (98.98-100.00%) on swabs and 100% (99.72-100.00%) on saliva when compared to RT-qPCR; analysing samples with RT-qPCR *ORF1ab* C_T_ values of ≤25 and ≤33, DSe of 100% (96.34-100%) and 77.78% (70.99-83.62%) for swabs were observed, and 99.01% (94.61-99.97%) and 87.61% (82.69-91.54%) for saliva, respectively. For RNA RT-LAMP, overall DSe and DSp were 96.06% (92.88-98.12%) and 99.99% (99.95-100%) for swabs, and 80.65% (73.54-86.54%) and 99.99% (99.95-100%) for saliva, respectively. These findings demonstrate that RT-LAMP is applicable to a variety of use-cases, including frequent, interval-based testing of saliva with Direct RT-LAMP from asymptomatic individuals that may otherwise be missed using symptomatic testing alone.

## Introduction

Rapid diagnostic testing to identify and isolate symptomatic and asymptomatic individuals potentially transmitting infectious viral pathogens is an essential requirement of any pandemic response. The novel *betacoronavirus*, SARS-CoV-2, initially identified after an outbreak of viral pneumonia in Wuhan, China in December 2019^1^, has rapidly spread throughout the world, causing over 223 million confirmed cases and over 4.6 million deaths (https://coronavirus.jhu.edu/ -September 10,2021).

Conventional diagnostics for SARS-CoV-2 consist of RNA enrichment followed by reverse-transcriptase quantitative real-time PCR (RT-qPCR) against one or more viral gene targets^2^. However, this methodology requires sample inactivation, RNA extraction and RT-qPCR thermal cycling, meaning that the time from sample-to-result can often be several hours, and requires centralised equipment and personnel trained in Good Laboratory Practice to perform testing.

The utility of reverse-transcriptase loop mediated isothermal amplification (RT-LAMP) for the detection of SARS-CoV-2 both from extracted RNA (RNA RT-LAMP) and directly from NP/OP swabs (Direct RT-LAMP)^3^ has previously been shown. RT-LAMP utilises a rapid and stable DNA polymerase that amplifies target nucleic acids at a constant temperature. This removes the requirement for conventional thermal cycling allowing RT-LAMP reactions to be performed in shorter reaction times using less sophisticated platforms.

In a study of 196 clinical samples^3^, testing of RNA extracted from NP/OP swabs collected into viral transport media (VTM) using RNA RT-LAMP demonstrated a diagnostic sensitivity (DSe) of 97% and a diagnostic specificity (DSp) of 99% in comparison to RT-qPCR of the ORF1ab region of SARS-CoV-2. For Direct RT-LAMP on crude swab samples, the DSe and DSp were 67% and 97%, respectively. When a cycle threshold (C_T_) cut-off for RT-qPCR of < 25 was considered, reflecting the increased likelihood of detecting viral RNA from active viral replication, the DSe of Direct RT-LAMP increased to 100% ^3^.

However, the collection of a swab sample is invasive and during the time of the pandemic there have been considerable shortages in swab supplies. Exploring the use of alternative sample types that are both easy to collect and more comfortable from a sampling perspective ^4,5,6^ is desirable particularly when repeat sampling is performed^7,8,9^. Saliva presents an ideal bio-fluid that fulfils both these objectives and previous studies have shown that SARS-CoV-2 is readily detectable in such a sample type^10,11,12–18^. To improve the diagnostic sensitivity of previously described saliva Direct RT-LAMP^3^, optimisation of saliva preparation for the detection of SARS-CoV-2 was undertaken utilising a cohort of 3100 saliva samples from an asymptomatic population^19^ of healthcare workers; where saliva was diluted 1:1 in Mucolyse™, followed by a 1 in 10 dilution in 10% (w/v) Chelex^©^ 100 Resin ending with a 98°C heat step prior to RT-LAMP which resulted in optimal sensitivity and specificity.

Despite the benefits of this optimisation, the protocol added additional steps and reagents which increased chance for user error and made the automation of the process more challenging. This study therefore aimed to investigate a simpler process using a novel reagent, RapiLyze (OptiGene Ltd, Camberley, UK), which is a sample dilution buffer, followed by a two-minute heat-step. This novel sample preparation method was evaluated in combination with Direct RT-LAMP using samples collected from symptomatic National Health Service (NHS) patients and symptomatic and asymptomatic healthcare staff.

## Methods

### Ethical statement

All relevant ethical guidelines have been followed, and any necessary IRB and/or ethics committee approvals have been obtained. National Research Ethics Service Committee West Midlands - South Birmingham 2002/201 Amendment Number 4. All necessary written participant consent has been obtained and the appropriate institutional forms have been archived.

### Testing sites

The OptiGene Ltd. SARS-CoV-2 RT-LAMP assay was evaluated in nine sites, comprising Basingstoke and North Hampshire Hospital & Royal Hampshire County Hospital, Hampshire Hospitals NHS Foundation Trust; University Hospital Southampton; Animal and Plant Health Agency/MRC Lifecourse Epidemiology Unit (University of Southampton); Public Health Lab Manchester/CMFT; Leeds Teaching Hospitals NHS Trust; University Hospitals Birmingham (UHB) NHS Foundation Trust/Institute of Cancer & Genomic Science University of Birmingham; High Containment Microbiology, National Infection Service, Public Health England, Porton Down and Public Health University Laboratory, Gibraltar Health Authority, Gibraltar, UK.

### Clinical samples

Nasopharyngeal (NP) and oropharyngeal (OP) swabs were collected from asymptomatic and symptomatic individuals and placed in viral transport media (VTM).

Drooled saliva samples were collected at the start of the day; prior to eating, drinking, teeth brushing, or using a mouthwash. Saliva was transferred into the specimen pot directly or via a clean teaspoon, according to a standardised protocol. Samples from UHB deposited saliva straight into the collection pot.

### Log reduction of SARS-CoV-2 for the heat and lysis steps used independently and sequentially

We determined the viral inactivation kinetics of the best sample preparation condition(s), evaluating the effect of the heat and lysis steps on the viral inactivation of SARS-CoV-2 as determined by infectivity assays. All inactivation experiments were conducted under Containment Level 3 Containment and as such were undertaken at APHA. Heat inactivation experiments were conducted utilising high titre live SARS-CoV-2 virus spiked into pools of saliva collected from APHA staff or in tissue culture supernatant (TCSN). Early experiments demonstrated that saliva had a high toxicity for tissue culture cells, even after heat inactivation demonstrating that toxicity was likely not enzymatic. As such further inactivation was undertaken on live virus TSCN. Comparison was also undertaken of Betapropiolactone (BPL) inactivated virus and live virus.

## RNA extraction

### RNA was extracted using a range of different methods available at each participating site

#### Maxwell® RSC Viral Total Nucleic Acid Purification Kit

In a class 1 microbiological safety cabinet (MSC) within a containment level 3 laboratory, 200 µl of sample was added to 223 µl of prepared lysis solution (including 5 µl per reaction of Genesig® Easy RNA Internal extraction control, Primerdesign Ltd, Chandler’s Ford, UK). Samples were then inactivated for 10 minutes at room temperature within the MSC and 10 minutes at 56°C on a heat block before automated RNA extraction using a Maxwell® RSC48 Instrument (Promega UK Ltd., Southampton, UK). RNA was eluted in 50 µl of nuclease-free water (NFW).

#### MagMAX™CORE Nucleic acid 140 purification kit

10 µl of sample (diluted in 190µl DEPC treated water) was added to 700 µl of prepared lysis solution. Samples were then inactivated for 10 minutes at room temperature within the safety cabinet before automated RNA extraction using a Kingfisher Flex (Thermo Fisher, Basingstoke, UK). RNA was eluted in 90 µl of NFW.

#### Roche FLOW system

RNA extraction was carried out on a MagNA Pure 96 (MP96) extraction robot using the MagNA Pure 96 DNA and Viral Nucleic Acid Small Volume kit (Roche, Basel, Switzerland) and the Pathogen 200 universal protocol v4.0.

#### Qiagen QIAsymphony

RNA extraction was carried out using the QIASymphony Virus/Bacteria Mini Kit (Qiagen, Hilden, Germany) by the CellFree200 Default IC protocol with a 60 µl extract elution volume.

### SARS-CoV-2 Real-Time RT-qPCR

RNA was analysed using a range of different methods available at each site:

#### CerTest VIASURE SARS-CoV-2 real time qPCR assay

Single step RT-qPCR against the *ORF1ab* region and *N1* gene target of SARS-CoV-2 was carried out using the CerTest VIASURE SARS-CoV-2 real time PCR kit (CerTest Biotech SL, Zaragoza, Spain) according to manufacturer’s instructions for use (IFU) on either the Thermo Fisher QuantStudio 5 or BioMolecular Systems (Queensland, Australia) MIC instruments, using 5 µl of extracted RNA per reaction. RNA extracted using the Maxwell® RSC Viral Total Nucleic Acid Purification Kit was analysed using this assay.

#### COVID-19 genesig® Real-Time qPCR assay

Single step RT-qPCR against the *ORF1ab* region of SARS-CoV-2 was carried out using the COVID-19 genesig® Real-Time PCR assay real time PCR assay (Primerdesign Ltd, Chandler’s Ford, UK) according to manufacturer’s instructions for use (IFU) on BioMolecular Systems MIC instruments, using 5 µl of extracted RNA per reaction. RNA extracted using the Maxwell® RSC Viral Total Nucleic Acid Purification Kit was analysed using this assay.

#### Corman et al. Real-Time qPCR assay

Single step RT-qPCR against the *E* gene target of SARS-CoV-2 was carried out with the Corman *et al*.^2^ primers using the AgPath-ID™ PCR kit (Thermo Fisher) according to manufacturer’s instructions for use (IFU) on an Aria qPCR Cycler (Agilent, Cheadle, UK) and results analysed using the Agilent AriaMX 1.5 software, using 5 µl of extracted RNA per reaction. RNA extracted using the MagMAX™CORE Nucleic acid purification kit were analysed using this assay.

RT-qPCR was carried out on an Applied Biosystems Fast 7500 PCR thermocycler in standard run mode using the SARS-CoV-2 *E* gene Sarbeco assay using MS2 as an internal extraction control and aliquots of SARS-CoV-2/England/2/2020 as a positive control. The master mix comprised *E*-gene F and R primers and TM-P (400 nM, 400 nM and 200 nM final concentration respectively), MS2 primers and TM probe (20 nM, 20 nM and 40 nM final concentration respectively), 4 x TaqMan® Fast Virus 1-Step Master Mix made up with molecular-grade nuclease free water (Ambion) to a final volume of 15 μl. 5 μl of AVE buffer extract was used at a template and added to the 15 μl mastermix. Cycling conditions were 55°C for 10 min, followed by 94°C for 3 min and 45 cycles of 95°C for 15 s and 58°C for 30 s.

#### SARS-CoV-2 (2019-nCoV) CDC qPCR Probe Assay

Single step RT-qPCR against the *N1* and *N2* gene targets of SARS-CoV-2 was carried out using integrated design technologies kit (IDT; Catalogue number: 10006606) according to manufacturer’s instructions for use (IFU) on either a LC480 II or ABI 7500 FAST instrument. RNA extracted on Qiagen QIAsymphony and the Roche FLOW system were analysed using this RT-qPCR assay.

### RT-LAMP

RT-LAMP assays were performed using OptiGene Ltd. COVID-19_RT-LAMP kits, as described previously^3^, with the following modifications. The COVID-19_RNA RT-LAMP KIT-500 kit was used for RNA RT-LAMP and the COVID-19_Direct PLUS RT-LAMP KIT-500 was used for Direct RT-LAMP directly on oropharyngeal/nasopharyngeal swabs or saliva samples. The COVID-19_Direct PLUS RT-LAMP KIT-500 kit also includes a sample preparation buffer, RapiLyze. For RNA RT-LAMP 5 μl of extracted RNA was added to the reaction. For the Direct PLUS RT-LAMP, 50 µl sample (swab VTM or neat saliva) was added to 50 µl RapiLyze, vortexed and placed in a dry heat block pre-heated to 98°C for 2 mins. 5 μl of the treated sample was added to each reaction.

The anneal temperature (Ta) that confirmed a positive result for Direct RT-LAMP was modified to 81.5°C and 85.99°C because of the effect of RapiLyze buffer on the reaction.

### SARS-CoV-2 viral culture of clinical samples across a C_T_ range

For culture, 100 µl and 100 µl of a 1 in 10 dilution of samples 1-6 (predicted lower C_T_ values) and 100 µl samples 7-26 (with higher predicted C_T_ values) were added to a 25 cm^2^ flasks containing 80% confluent Vero E6 cells and allowed to adsorb for 1 hour. Five ml of Minimum Essential Medium (MEM) (Gibco) + HEPES (Gibco, Thermo Fisher, Basingstoke, UK) + 4% foetal calf serum FCS (Sigma) + 1 x antibiotic-antimycotic (Gibco) was added to each flask and incubated for 1 week at 37°C. Two negative control flasks to which 100 µl MEM + 4% FCS was added in place of sample, were set up in parallel. Cultures were checked visually for cytopathic effect (CPE). Where CPE was not observed after 1 week, 500 µl of supernatant was passed into a fresh flask containing Vero E6 cells for a further two passages. At the beginning and end of each passage 140 µl of supernatant was collected for SARS-CoV-2 RT-qPCR as described before.

To determine the sensitivity of the isolation method for SARS-CoV-2 from clinical samples, a virus stock titred by plaque assay (HCM/V/53), a passage 3 working bank grown from SARS-CoV-2 Strain England 2, from Public Health England, was diluted in MEM to give virus dilutions containing 1000, 100, 10, 1, 0.1 and 0.01 PFU. The virus was added to duplicate flasks containing Vero E6 cells and AVL. After 72 hours of incubation flasks were checked for CPE, and for those where CPE was observed the supernatant was collected for RT-qPCR. Any flasks not showing CPE after 7 days were passed on to fresh cells and resampled as described above.

## Data analysis

Overall diagnostic sensitivity and specificity (including 95% Clopper-Pearson confidence intervals) were calculated by the aggregation of individual site data for each method (RNA and Direct RT-LAMP) for each sample type (swabs and saliva). To demonstrate the effectiveness of detecting samples with higher viral load, confusion matrices are quoted where the threshold for *positive* sample inclusion varies, i.e., for C_T_ ≤25, only positive samples with C_T_ ≤25 are included.

To account for site heterogeneity, a bivariate meta-analysis model is additionally applied at the site level to produce a summary of sensitivity and specificity for each method and sample type^20^. Within-study variability for sensitivity *ρ*_*se,i*_ and specificity *ρ*_*se,i*_ are assumed to follow independent binomial

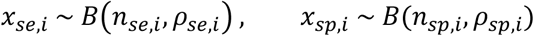

distributions where *x*_*se,i*,_*x*_*sp,i*_ represent the number testing positive for site *i* respectively, and *n*_*se,i*,_*n*_*sp,i*_ represent the number testing positive and negative by RT-qPCR for site *i* respectively. The between-study heterogeneity is represented by a bivariate normal distribution for the logit-transformed sensitivity *µ*_*se,i*_ and specificity *µ*_*sp*.*i*_

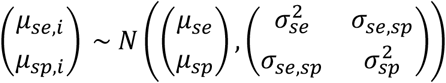

where *µ*_*se*,_*µ*_*sp*_ represent the expected logit sensitivity and specificity, 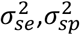 represent the between-study variance in the logit sensitivity and specificity, and *σ*_*se,sp*_ represents the covariance between the logit sensitivity and specificity. For Direct RT-LAMP, we fit a univariate normal distribution for the logit-transformed sensitivity only, due to the absence of false positives across all sites.

In addition, the sensitivity as a function of viral load was assessed for RNA RT-LAMP and Direct RT-LAMP on both swab and saliva samples. This was performed through the conversion of each sample C_T_ value to viral load in gene copies/ml for all sample sets. As the relationship between C_T_ value and viral load varied according to the RT-qPCR method used; a dilution series was utilised for each method to standardise these values for two of the four aforementioned RT-qPCR methods (CerTest VIASURE SARS-CoV-2 real time PCR kit, and Corman *et al* RT-qPCR assay *E* gene), which was used for testing 100% of the swab samples, 90% of the saliva samples used for Direct RT-LAMP, and 83% of the saliva samples used for RNA RT-LAMP. The logarithm of the viral load was then fitted to the C_T_ values for both methods using linear regression followed by converting the C_T_ values to viral load based on which method had been used to evaluate the samples. For the remaining samples (n= 56) that utilised one of the other two RT-qPCR methods for which viral load was not standardised against a C_T_ value, the conversion derived from the CerTest VIASURE SARS-CoV-2 real time PCR kit dilution series was applied, the assumption that the *N* gene C_T_ values are the most similar^21–23^.

For the CerTest VIASURE SARS-CoV-2 real time PCR kit, the following relationship between log viral load and C_T_ value was applied:

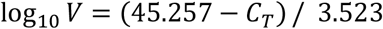

and similarly, for the Corman *et al* RT-qPCR assay:

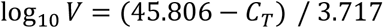

where *V* represents the viral load in copies/ml.

Viral load was grouped according to the following categories (in copies/ml): <10^3^, 10^3^-10^4^, 10^4^-10^5^, 10^5^- 10^6^, 10^6^-10^7^ and >10^7^ then the diagnostic sensitivity was calculated according to viral load group with associated Clopper-Pearson 95% confidence intervals.

The site meta-analysis was produced using R 3.5.3. Confusion matrices, sensitivity, specificity, sensitivity as a function of viral load calculations, and the production of scatter graphs showing the relationship between RT-LAMP results and C_T_ were performed using Python 3.8.6.

## Results

### Optimisation of sample preparation conditions

Heat inactivation experiments demonstrated that SARS-CoV-2 was completely inactivated by heating at 60°C (20 min plus) or ≥70°C (after 2, 5 or 10 min) (Supplemental Table S1). Importantly optimised RapiLyze Sample Lysis Buffer did not inactivate live virus on its own without a heat step. Further, inactivation at 56°C was not 100% effective at shorter incubation times, and additionally showed a loss in sensitivity following a 4 × 2-fold dilution (Supplemental Table S2, P07102) at 10 and 30 minutes. Following optimisation of heat inactivation of live virus, pre-treatment of samples was assessed to determine any impact of pre-treatment on assay sensitivity. Interestingly, a pre-treatment 70°C for 5 mins carried out on spiked samples prior to the proposed direct RT-LAMP assay had no effect on subsequent LAMP or PCR results. It recommended that even if a pre-treatment is effective in inactivating the virus that downstream processes are carried out in UV hoods or with effective air-flow management to prevent cross contamination of the direct RT-LAMP assay. Comparison of Betapropiolactone (BPL) inactivated virus and live virus demonstrated that BPL inactivation has resulted in lower sensitivity of detection using direct RT-LAMP. BPL inactivated virus is therefore not an ideal substitute for live virus in spiking experiments. Any conclusions on assay sensitivity or performance have consequently been drawn from experiments on spiking of live virus in TCSN or saliva carried out in containment. Spiking of live virus into pooled saliva has demonstrated that direct detection by RT-LAMP is possible in samples that give a C_T_ below 25/26 with extraction and PCR.

### RNA RT-LAMP

VTM from 12,619 NP/OP swabs were assayed. 265 swab samples were from known symptomatic individuals and 2073 swab samples were from known asymptomatic individuals. The clinical status of the remaining samples (n= 10,281) was unknown.

12,521 neat saliva samples were assayed, none of which were from known symptomatic individuals. 12,365 of these samples were from known asymptomatic individuals. The clinical status of the remaining saliva samples (n= 156) was unknown.

### Direct RT-LAMP

VTM from 559 NP/OP swabs were assayed. 170 swab samples were from known symptomatic individuals and 310 samples were from known asymptomatic individuals and the clinical status of the remaining swab samples (n= 79) was unknown.

86,760 neat saliva samples were assayed. 93 samples were from known symptomatic individuals and 86,593 samples were from known asymptomatic individuals. The clinical status of the remaining samples (n= 74) was unknown. In addition, 10 separate longitudinal daily saliva samples were provided from one individual as a time course from development of symptoms to three days post resolution of symptoms.

### RNA RT-LAMP on NP/OP swabs

A total of 12,619 swab samples were assayed by RNA RT-LAMP, of which 254 were RT-qPCR positive and 12,365 were RT-qPCR negative. RNA RT-LAMP detected 244 of the 254 positives (Figure 1 and Table 1). Only one of the 12,365 samples negative by RT-qPCR was positive by RNA RT-LAMP. 588 samples were tested in duplicate and 12,031 were tested as single replicates. Of those samples tested in duplicate seven were detected by RNA RT-LAMP in only a single replicate (C_T_s 27.00, 32.66, 33.14, 33.16, 34.07, 35.05, and 37.20 all of these had received at least one freeze thaw before analysis. Overall diagnostic sensitivity (DSe) was 96.06% (95% CI 92.88-98.12) and specificity (DSp) 99.99% (95% CI 99.95-100.00), which is corrected to DSe 95.98% (95% CI 92.70-97.83) and DSp 99.99% (95% CI 99.94-100.00) after site meta-analysis. Diagnostic sensitivity of samples with a C_T_ <25 (n= 123) was 100.00% (95% CI 96.76-100.00) and specificity 99.99% (95% CI 99.95-100.00), and of samples with a C_T_ <33 (n=180) was 98.65% (95% CI 96.10-99.72) and specificity 99.99% (95% CI 99.95-100.00).

**Figure 1.**
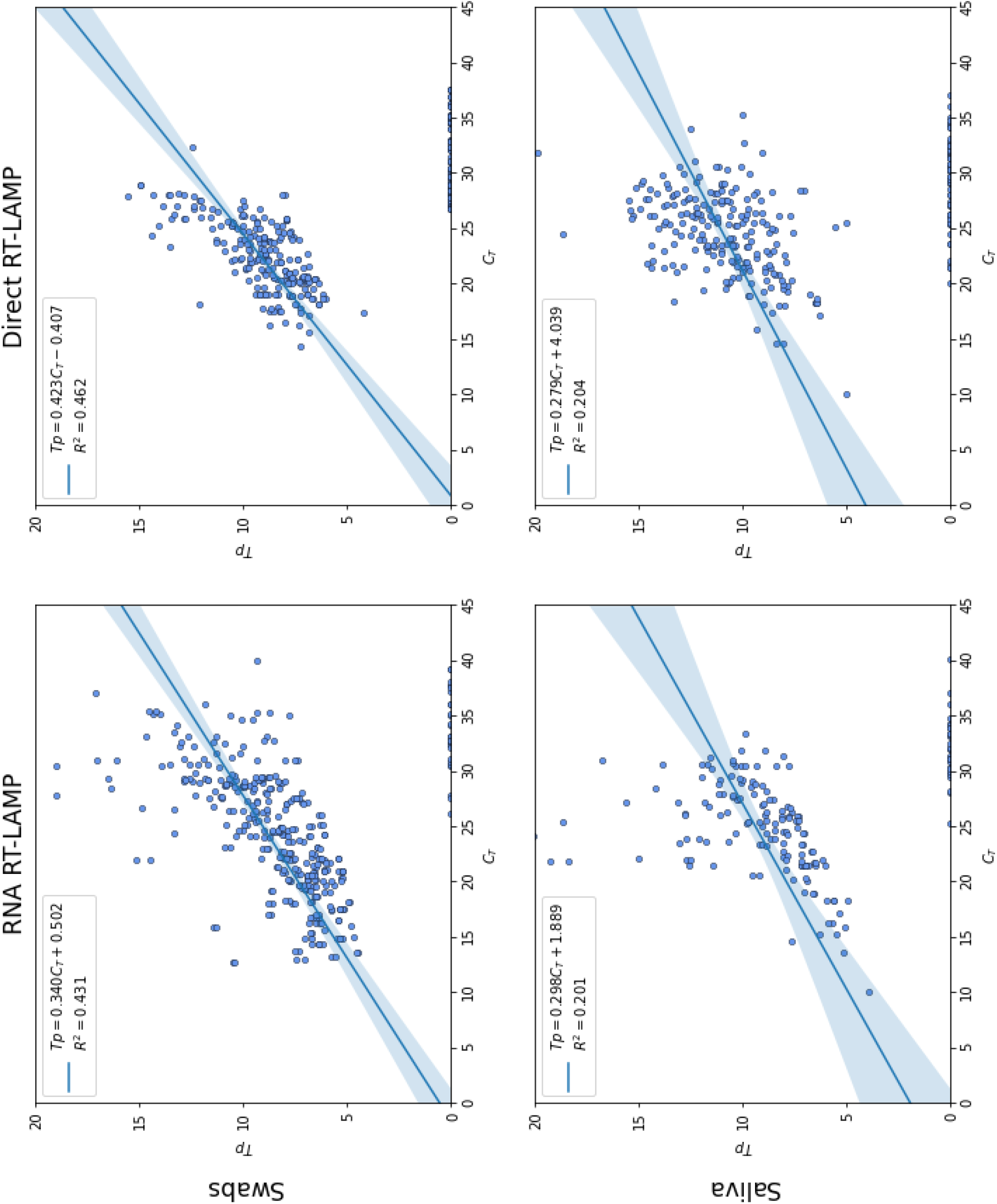
Time to positivity [Tp] in minutes plotted against RT-qPCR Cycle Threshold [C_T_] for each combination of method and sample type. Samples which were negative by RT-qPCR are not shown. Samples which were negative by RT-LAMP are shown with 0 time to positivity. Results of linear ordinary least squared regression are shown for samples which were RT-LAMP positive with the regression line and corresponding 95% confidence interval represented by the blue line and light blue shaded regions respectively.

**Table 1.** Diagnostic sensitivity and specificity of RNA RT-LAMP on swabs compared with RT-qPCR

|  |  | RT-qPCR Pos | RT-qPCR Neg | Total |  | % | 95% CI |
| --- | --- | --- | --- | --- | --- | --- | --- |
| **C<sub>T</sub> <45 Swab** | RNA RT-LAMP Pos | 244<sup>†</sup> | 1 | 245 | **DSe** | 96.06 | 92.88-98.12 |
|  | RNA RT-LAMP Neg | 10 | 12364 | 12374 | **DSp** | 99.99 | 99.95-100 |
|  | Total | 254 | 12365 |  |  |  |  |
|  |  | RT-qPCR Pos | RT-qPCR Neg | Total |  | % | 95% CI |
| **C<sub>T</sub> <33 Swab** | RNA RT-LAMP Pos | 219 | 1 | 220 | **DSe** | 98.95 | 96.10-99.72 |
|  | RNA RT-LAMP Neg | 3 | 12364 | 12367 | **DSp** | 99.9 | 99.95-100 |
|  | Total | 222 | 12365 |  |  |  |  |
|  |  | RT-qPCR Pos | RT-qPCR Neg | Total |  | % | 95% CI |
| **C<sub>T</sub> <25 Swab** | RNA RT-LAMP Pos | 112 | 1 | 113 | **DSe** | 100 | 96.76-100 |
|  | RNA RT-LAMP Neg | 0 | 12364 | 12364 | **DSp** | 99.9 | 99.95-100 |
|  | Total | 112 | 12365 |  |  |  |  |

<sup>†</sup>Five samples included in this number were positive by RT-qPCR but did not have an associated C<sub>T</sub> value due to being assayed on a platform that did not produce a C<sub>T</sub> value. DSe: Diagnostic sensitivity. DSp: Diagnostic specificity

### Direct RT-LAMP on NP/OP swabs

559 swab samples were assayed by Direct RT-LAMP of which 199 were RT-qPCR positive and 360 were RT-qPCR negative. Direct RT-LAMP detected 140 of the 199 samples positive by RT-qPCR (Figure 1 and Table 2). 195 samples were tested in duplicate and 364 tested as single replicates. Seven of 195 samples tested in duplicate were positive by Direct RT-LAMP in only one replicate (C_T_ 27.51, 27.95, 28.15, 28.15, 28.87, 28.92, and 28.95) all these samples had received at least one freeze thaw before analysis. Overall diagnostic sensitivity was 70.35% (95% CI 63.48-76.60) and specificity 100% (95% CI 98.98-100). After correction by site meta-analysis, the DSe is corrected to 67.59% (95% CI 53.71-78.94). Diagnostic sensitivity of samples with a C_T_ <25 (n= 113) was 100% (95% CI96.34-100) and specificity 100% (95% CI 98.98-100), and of samples with a C_T_ <33 (n= 182) was 77.78% (95% CI 70.99-83.62) and specificity 100% (95% CI 98.98-100).

**Table 2.** Diagnostic sensitivity and specificity of Direct RT-LAMP on swabs compared to RT-qPCR

|  |  | RT-qPCR Pos | RT-qPCR Neg | Total |  | % | 95% CI |
| --- | --- | --- | --- | --- | --- | --- | --- |
| **C<sub>T</sub> <45 Swab** | Direct RT-LAMP Pos | 140 | 0 | 140 | **DSe** | 70.35 | 63.48-76.60 |
|  | Direct RT-LAMP Neg | 59 | 360 | 419 | **DSp** | 100 | 98.98-100 |
|  | Total | 199 | 360 |  |  |  |  |
|  |  | RT-qPCR Pos | RT-qPCR Neg | Total |  | % | 95% CI |
| **C<sub>T</sub> <33 Swab** | Direct RT-LAMP Pos | 140 | 0 | 140 | **DSe** | 77.78 | 70.99-83.62 |
|  | Direct RT-LAMP Neg | 40 | 360 | 400 | **DSp** | 100 | 98.98-100 |
|  | Total | 180 | 360 |  |  |  |  |
|  |  | RT-qPCR Pos | RT-qPCR Neg | Total |  | % | 95% CI |
| **C<sub>T</sub> <25 Swab** | Direct RT-LAMP Pos | 99 | 0 | 99 | **DSe** | 100 | 96.34-100 |
|  | Direct RT-LAMP Neg | 0 | 360 | 360 | **DSp** | 100 | 98.98-100 |
|  | Total | 99 | 360 |  |  |  |  |

DSe: Diagnostic sensitivity. DSp: Diagnostic specificity

### RNA RT-LAMP on saliva

Saliva samples numbering 12,521 were assayed by RNA RT-LAMP of which 155 were RT-qPCR positive and 12,366 were RT-qPCR negative. RNA RT-LAMP detected 133 of the 155 samples that were positive by RT-qPCR (Figure 1 and Table 3). Only one of the 12,366 samples negative by RT-qPCR was positive by RNA RT-LAMP. 44 samples were tested in duplicate and 12,477 were tested as single replicates. All samples tested in duplicate were positive in both replicates. Overall diagnostic sensitivity was 80.65% (95% CI 73.54-86.54) and specificity 99.99% (95% CI 99.95-100), which is corrected to DSe 79.05% (95% CI 68.87 – 86.55) and DSp 99.99% (95% CI 99.74-100) after site meta-analysis. Diagnostic sensitivity of samples with a C_T_ <25 (n= 74) was 100% (95% CI 93.73-100) and specificity 99.99% (95% CI 99.95-100), and of samples with a C_T_ <33 (n= 150) was 87.32% (95% CI 80.71-92.31) and specificity 99.95 (95% CI 99.95-100.00).

**Table 3.**
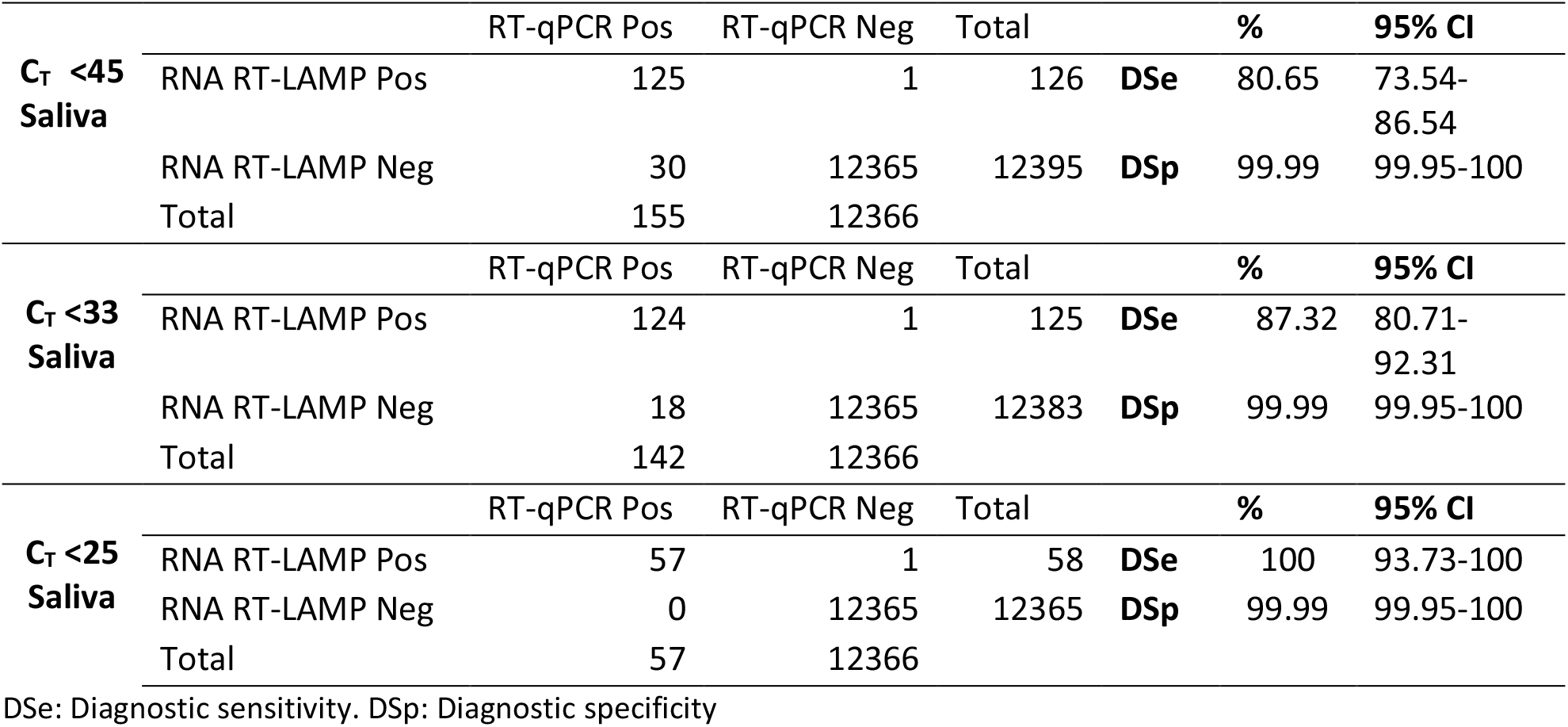
Diagnostic sensitivity and specificity of RNA RT-LAMP on saliva compared to RT-qPCR

### Direct RT-LAMP on saliva

86,760 saliva samples were tested by Direct RT-LAMP of which 247 were RT-qPCR positive and 7,195 were RT-qPCR negative (79,318 were negative on RT-LAMP but were not tested by RT-qPCR) (Figure 1 and Table 4). Direct RT-LAMP detected 209 of the 247 samples positive by RT-qPCR. 83 samples were tested in duplicate and 86,677 were tested as single replicates. Nine of the 83 samples tested in duplicate were negative in one of the duplicates and all these samples had received at least one freeze thaw before analysis (C_T_ 20.27, 21.28, 22.01, 24.42, 25.85, 27.35, 28.52, and 30.37). Overall diagnostic sensitivity was 84.62% (95% CI 79.50-88.88) and specificity 100% (95% CI 99.72-100). After correction by site meta-analysis, DSe is corrected to 84.24% (95% CI 55.03-95.89). Diagnostic specificity was calculated using only the confirmed RT-qPCR negative samples. Diagnostic sensitivity of samples with a C_T_ <25 (n= 126) was 99.01% (95% CI 94.61-99.97) and specificity 100.00% (95% CI 99.72-100), and of samples with a C_T_ <33 (n= 237) was 87.61% (95% CI 82.69-91.54) and specificity 100% (95% CI 99.72-100).

**Table 4.**
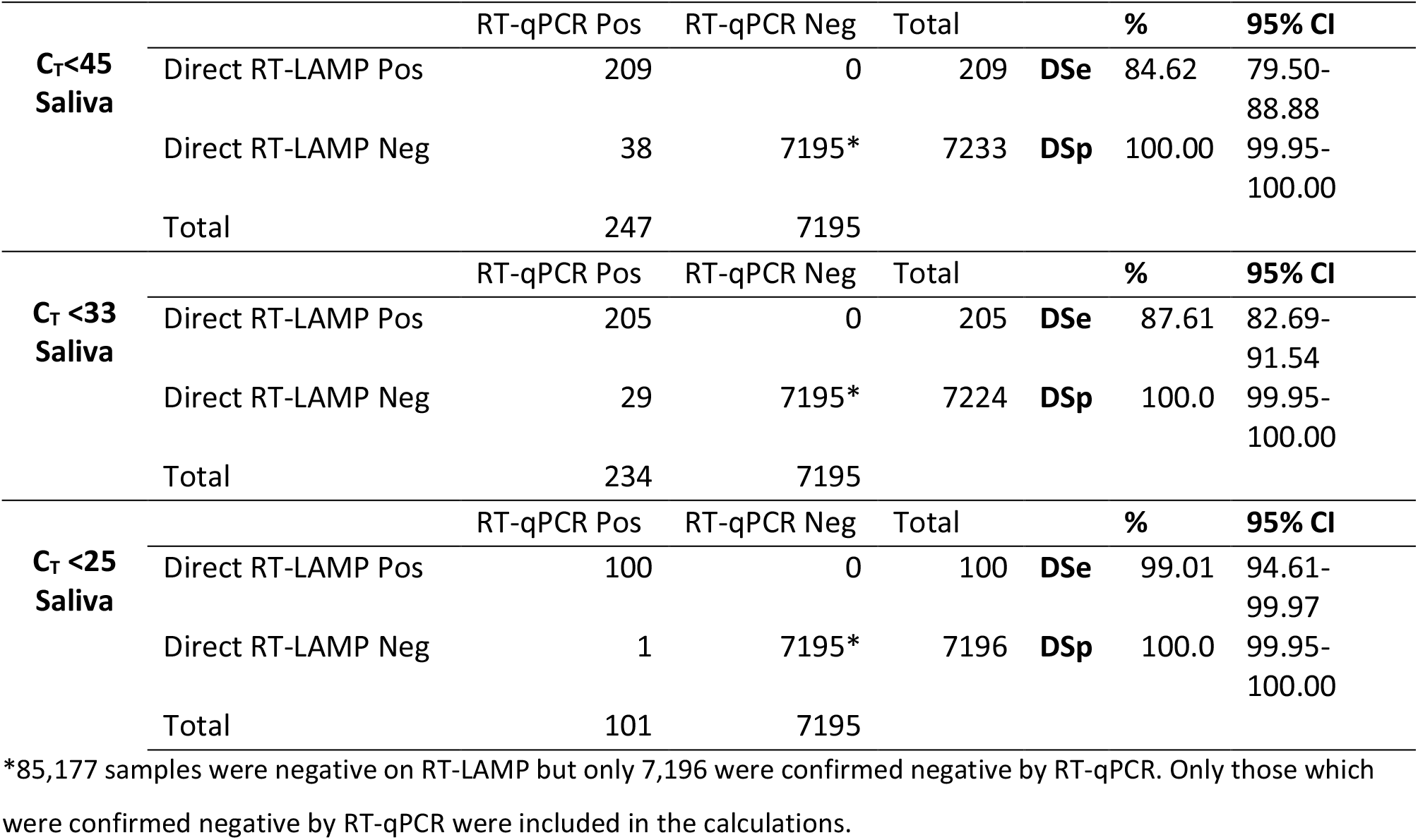
Diagnostic sensitivity and specificity of Direct RT-LAMP on saliva compared to RT-qPCR.

### Relationship between cycle threshold (C_T_) value and time to positivity (Tp)

The relationship between C_T_ value and Tp was explored with the results shown in Figure 1. Whilst there is a weak linear relationship between C_T_ value and Tp across all methods and sample types, a stronger linear relationship was observed in swab samples with *R*^2^ = 0.431 for RNA RT-LAMP and *R*^2^ = 0.462 for Direct RT-LAMP. There was a notably weaker linear relationship in saliva samples *R*^2^ = 0.201 for RNA RT-LAMP and *R*^2^ = 0.204 for Direct RT-LAMP. For RNA RT-LAMP, there was a notable increase in Tp variance, 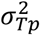, after C_T_ = 20 across both sample types. On saliva samples, 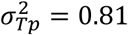 for C_T_≤20, and 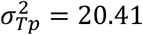 for C_T_ >20; on swabs samples, 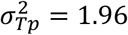 and C_T_ <20, and 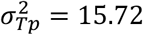 for C_T_ >20. Given the relationship between C_T_ value and viral load, this indicates that Tp is not a reliable indicator for viral load beyond the C_T_ = 20 threshold.

### SARS-CoV-2 viral culture of clinical samples across a C_T_ range

Although not a large sample size; a negative result via Direct RT-LAMP indicates that the presence of culturable virus is less probable and for samples with a C_T_ >25 (RDRP/ORF1ab target) recoverable virus is less likely (Table 5). The sensitivity of 1 PFU/ml of the viral culture assay is presented in Supplemental Table S3. No CPE was observed in the flasks inoculated with 0.1 or 0.01 PFU after the two passes. AVL samples were taken from the flasks at the beginning and end of each passage and the C_T_ values of the extracted nucleic acids shown in Supplemental Table S3.

**Table 5.** Viral culture of positive VTM from oro/pharyngeal swabs and assay results.

| Sample | Direct RT-LAMP | RNA RT-LAMP | C<sub>T</sub> values for each RT-qPCR assay |  |  | CPE |
| --- | --- | --- | --- | --- | --- | --- |
|  |  |  | Genesig RDRP gene | VIASURE | SARS-CoV-2 E gene Sarbeco assay |  |
|  |  |  |  | ORF1ab |  |  |
| 1 | POS | POS | 19.9 | 18.7 | 17.8 | CPE+ |
| 2 | POS | POS | 21.3 | 19.9 | 19.0 | CPE+ |
| 3 | POS | POS | 21.6 | 19.1 | 18.5 | CPE+ |
| 4 | POS | POS | 22.6 | 20.8 | 19.8 | CPE+ |
| 5 | POS | POS | 22.9 | 21.6 | 21.0 | CPE+ |
| 6 | POS | POS | 23.7 | 20.6 | 20.6 | CPE+ |
| 7 | NEG | POS | - | ND | ND | No CPE |
| 8 | NEG | POS | 39.2 | ND | ND | No CPE |
| 9 | NEG | POS | 35.2 | ND | ND | No CPE |
| 10 | NEG | NEG | 34.6 | ND | ND | No CPE |
| 11 | NEG | POS | 35.4 | ND | ND | No CPE |
| 12 | NEG | POS | 36.2 | ND | ND | No CPE |
| 13 | POS | POS | 35.8 | ND | ND | No CPE |
| 14 | POS | POS | 34.5 | ND | ND | No CPE |
| 15 | NEG | POS | 35.1 | ND | ND | No CPE |
| 16 | POS | POS | 30.0 | ND | ND | No CPE |
| 17 | POS | POS | 32.3 | ND | ND | No CPE |
| 18 | NEG | POS | 34.6 | ND | ND | No CPE |
| 19 | POS | POS | 31.3 | ND | ND | No CPE |
| 20 | NEG | POS | 30.3 | ND | ND | No CPE |
| 21 | NEG | POS | 30.0 | ND | ND | No CPE |
| 22 | NEG | POS | 31.5 | ND | ND | No CPE |
| 23 | NEG | POS | 30.7 | ND | ND | CPE+ |
| 24 | POS | POS | 29.9 | ND | ND | No CPE |
| 25 | POS | POS | 29.4 | ND | ND | No CPE |
| 26 | NEG | POS | 28.2 | ND | ND | CPE+ |

Samples were taken through 3 passages. ND: not done.

### Individual time course

In the time course experiment SARS-CoV-2 RNA was detected from day 5 (at the onset of symptoms) up to day 12 post suspected initial exposure using Direct RT-LAMP and up to day 13 by RNA RT-LAMP, encompassing the full six days where symptoms were recorded, Supplemental Table S4.

### Performance of RT-LAMP across viral load groups

The sensitivity of the RNA and Direct RT-LAMP assays across viral load groups is shown in Figure 2. For RNA RT-LAMP, samples which were positive by RT-qPCR containing >10^5^ copies/ml were consistently identified as positive with no samples returning a negative result. Below this copy number, sensitivity is reduced for both saliva and NP/OP swab samples, reaching ∼60% in NP/OP swab samples exclusively with viral loads <10^3^ copies/ml, and an approximately linear drop in sensitivity from 100% to 0% between viral loads of 10^5^ and 10^3^ copies/ml respectively in saliva samples. For Direct RT-LAMP, all but one saliva sample were detected above 10^6^ copies/ml. On swab samples, sensitivity is reduced on samples containing below <10^5^ copies/ml, dropping from 85% at viral loads of 10^5^–10^6^ copies/ml, to 30% in the 10^4^–10^5^range. On saliva samples, sensitivity is reduced in the 10^4^–10^5^ range to a sensitivity of 80% but then reduces further within the 10^3^–10^4^ range, to 30%.

**Figure 2.**
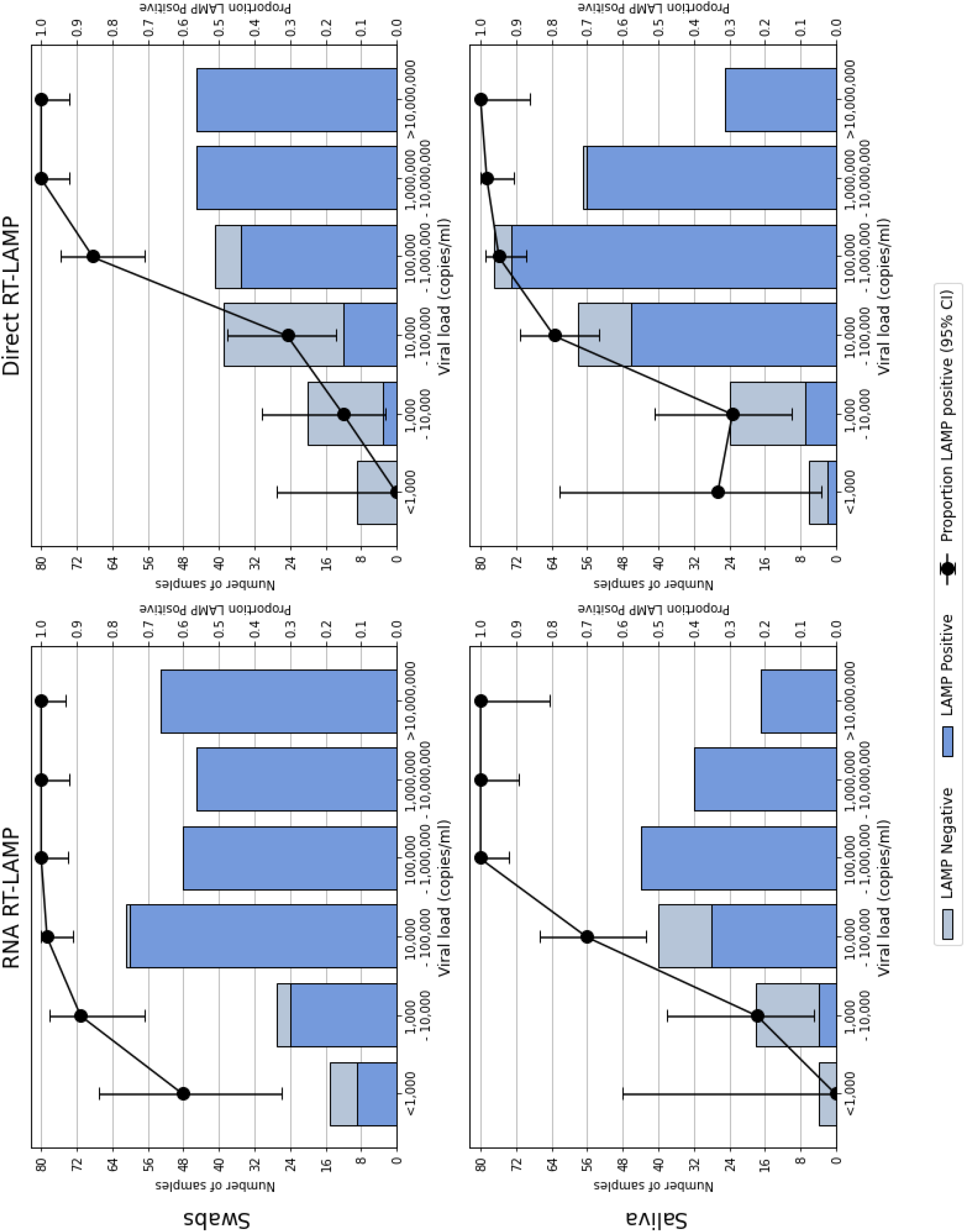
Performance of the RNA RT-LAMP and Direct RT-LAMP assays on both saliva and swab samples according to viral load groupings.

### Site meta-analysis

Site-level confusion matrices, sensitivity, and specificity per method and sample type are shown in Figures 3 and 4. For specificity, heterogeneity between sites was minimal for all combinations of method and sample type, with the random effects model matching the overall aggregated sample calculation. For sensitivity, heterogeneity was minimal between sites for RNA RT-LAMP. However, for Direct RT-LAMP, sensitivity showed significant overall heterogeneity (bivariate model variance: 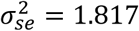 on saliva samples; 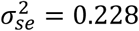 on swab samples). Between-site variations in the viral load of the samples tested contributed a minority of the heterogeneity, but sensitivity was consistently high in samples with higher viral loads (i.e., >10^6^ copies/ml, as shown in Figure 2), while being more heterogeneous between sites in samples with lower viral loads. Sensitivity at lower viral loads was highest in the sites with the most established testing programmes.

**Figure 3.**
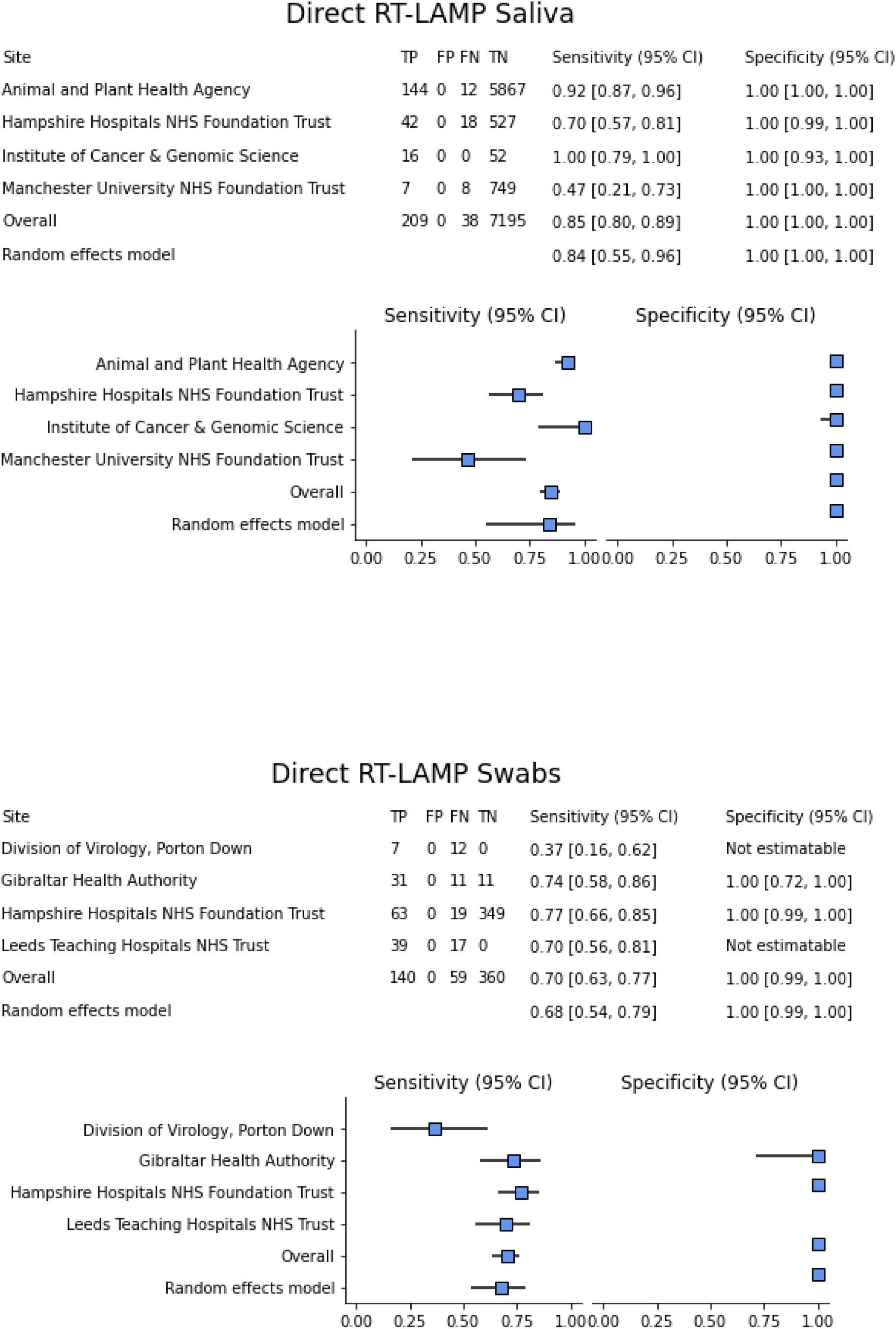
Forest plots for Direct RT-LAMP per sample type showing site heterogeneity in sensitivity and specificity, with overall estimates and the resulting expected sensitivity and specificity retrieved from each respective bivariate random effects model.

**Figure 4:**
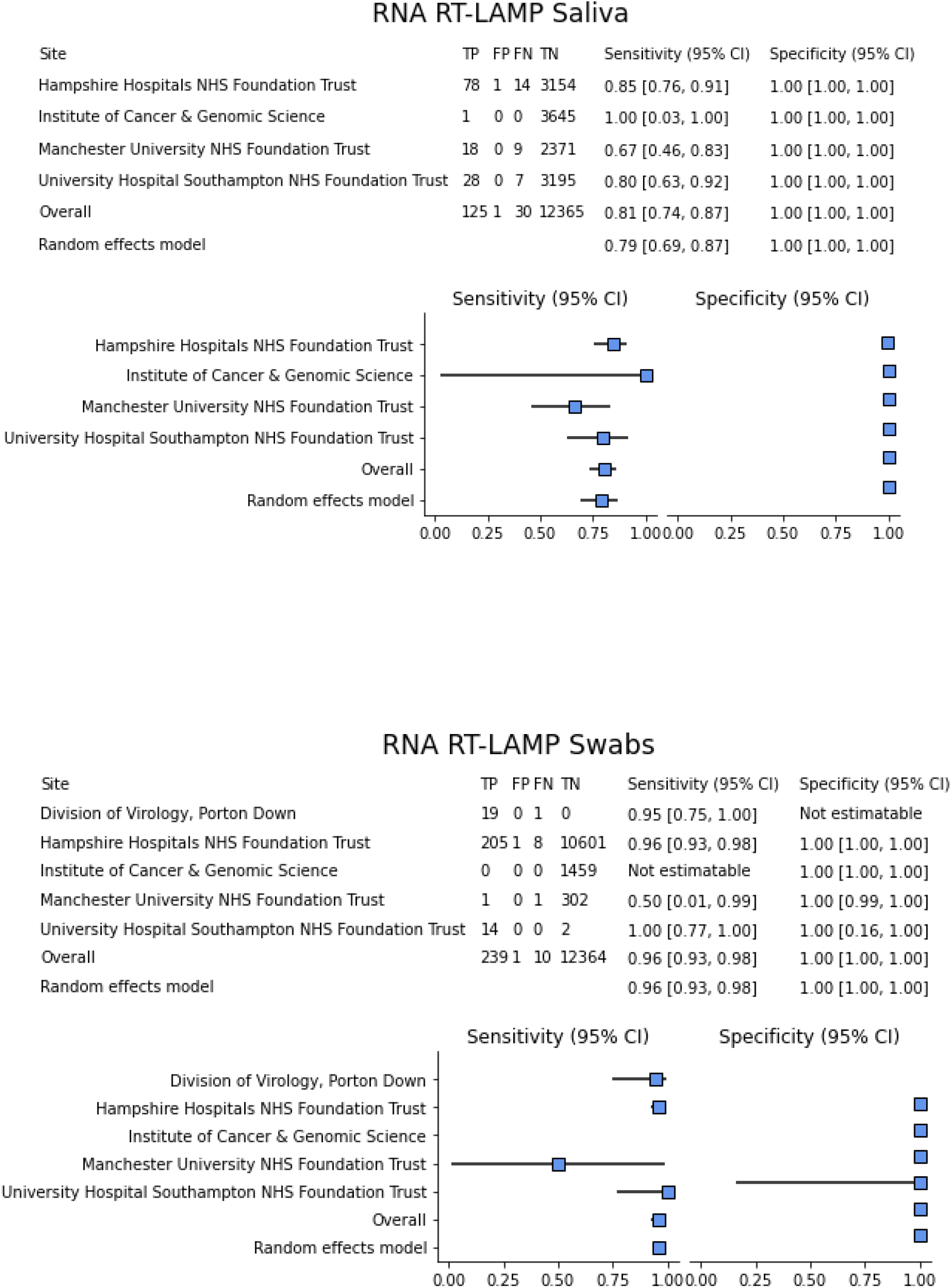
Forest plots for RNA RT-LAMP per sample type showing site heterogeneity in sensitivity and specificity, with overall estimates and the resulting expected sensitivity and specificity retrieved from each respective bivariate random effects model.

## Discussion

Testing of human populations for SARS-CoV-2 nucleic acid has been hampered by both logistical (e.g., swab availability) and physical (e.g., discomfort from repeat swab testing) constraints. The aim of this study was to evaluate an optimised sample preparation method, building upon previously published methods for the extraction-free detection of SARS-CoV-2 by RT-LAMP primarily from saliva^3,19^. The salivary glands are reported to be early targets of SARS-CoV-2 infection^24^, and studies have demonstrated the detection of high viral loads of SARS-CoV-2 from saliva, similar to those observed from nasopharyngeal/oropharyngeal swabs^15,25–27^. Collection of saliva is non-invasive and does not require a trained individual or specialist consumables for collection of a quality sample. Utilising a non-invasive sampling method should open testing to more individuals who dislike or are unable to tolerate having a nasopharyngeal/oropharyngeal swab taken^28^. Additionally, several studies have demonstrated that SARS-CoV-2 viral RNA could be detected from saliva for a similar duration post onset of clinical signs when compared to combined NP/OP swabs^29–31^, highlighting saliva as a valuable tool for SARS-CoV-2 detection.

Direct detection negates the requirement for RNA extraction^32,33^, for which there has previously been competition for reagents and often requires expensive extraction equipment including liquid handling automation. This extraction-free method decreases turnaround time from sample collection to result. The Direct RT-LAMP method is straight forward and rapid, allowing the test to be performed in a wide range of settings, including near-patient hospital laboratories and pop-up or mobile laboratories. However, previously evaluated extraction-free sample preparation methods using RT-LAMP from saliva samples have demonstrated reduced sensitivity^3,19^, likely due to the inhibitory factors found within saliva that may affect molecular tests such as RT-LAMP and RT-qPCR^34,35^. The simple sample preparation method evaluated in the study aimed to improve upon these methods by utilising the addition of a novel proprietary reagent, RapiLyze^©^, designed to neutralize common sample inhibitors. A subsequent heat step of 98°C for two minutes prior to addition to the RT-LAMP master mix renders SARS-CoV-2 inactive as confirmed by infectivity analysis using live virus inactivation studies (Supplemental Tables S1 and S2). Downstream steps are then able to proceed outside of traditional containment level laboratory settings broadening its clinical utility.

This study utilised high numbers of combined naso/oropharyngeal swabs (n= 559) and saliva samples (n = 86,760) for the evaluation of this novel sample preparation method in combination with the Direct RT-LAMP assay. RNA RT-LAMP was also performed on >25,000 samples for comparison, providing updated values for the performance of the assay reported previously^3,19,36^. Correlation between C_T_ value and sample viral copy number has been demonstrated within this and other studies, with lower C_T_ values (C_T_ <25 and <33) indicating a higher probability that the sample contains recoverable active virus, and consequently the likelihood that the individual may be infectious to others^4,25,37–40^. As a result, the RNA and Direct RT-LAMP assays were compared with RT-qPCR results in groups with three different C_T_ cut-off values: <45, <33 and <25. This was completed so that the performance of the assays in different clinical scenarios (use case) could be determined: (i) C_T_ <45: does the RT-LAMP assay (either RNA or Direct) compare with RT-qPCR for all reportable C_T_ values?; (ii) C_T_ <33: can the RT-LAMP assay detect those individuals that have a medium-high levels of viral RNA in their specimens, with an *ORF1ab* target being analogous with viral copy number because it is exclusively a genomic target^22^; and (iii) C_T_ <25: can the RT-LAMP assay detect those individuals that have a high level of viral RNA in their specimens?

Diagnostic sensitivity for RNA RT-LAMP on swab and saliva samples was improved when compared to a previous report utilising this method^3^, with values of >96% and >80%, respectively when considering all C_T_ values, and 100% for both sample types when considering C_T_ <25 with these samples having a high probability of containing replicating virus for over 24,000 samples tested. Direct RT-LAMP sensitivity on swab samples was also improved from the previous method with 100% sensitivity for C_T_ <25, 77.78% for C_T_ <33 and 70.35% for C_T_ <45 across 559 samples used for this evaluation. In contrast, sensitivity for Direct RT-LAMP on saliva was in general higher than that determined for swabs (C_T_ <33 = 87.61%, C_T_ <45 = 84.62%), apart from the group with C_T_ values below <25, which had a reported sensitivity of 99.01%. These results support previous reports which demonstrate comparable performance when comparing paired swabs and saliva samples^41,42^, and that one sample type is not superior to the other. Interestingly, the diagnostic sensitivity for RNA and Direct RT-LAMP for saliva samples was almost equivalent (80.65% and 84.62%, respectively) suggesting that RNA extraction may not even be required when performing testing on saliva samples. Direct RT-LAMP also demonstrates a higher sensitivity than a wide variety of lateral flow tests (LFTs) in the C_T_ < 25, C_T_ ≥ 25 and overall categories, with the overall sensitivity of Direct RT-LAMP on saliva samples achieving a higher overall sensitivity than 94 out of 96 LFTs previously evaluated^43^. We found that the correlation between PCR C_T_ value and the Direct RT-LAMP Tp was weaker for saliva than for swabs, which may reflect the PCR C_T_ value being from a naso-pharyngeal swab and recognised time course differences between initial viral infection of the salivary glands and later infection of the respiratory tract^26,30^.

Previous studies have described the importance of identifying asymptomatic individuals, particularly those with high viral loads^28,44–48^. The ability of the Direct RT-LAMP assay to reliably detect individuals with medium-high viral loads in a simple to collect, non-invasive sampling process highlights the suitability of this assay for both symptomatic and asymptomatic population screening. This is particularly important in healthcare and care home staff where the use of asymptomatic COVID-19 screening would reduce the risk of onward transmission of SARS-CoV-2, consequently maintaining NHS capacity and Social Care capacity and more importantly, reducing the risk to vulnerable individuals present within those environments^36^.

It is important to note that when designing surveillance strategies for asymptomatic infection testing as an intervention to reduce transmission, frequency of testing and result turnaround time may be considered more significant than diagnostic sensitivity^49^. ‘Gold standard’ tests with high sensitivity such as RT-qPCR generally need to be performed in centralised testing facilities, often resulting in increased reporting times, leading to a less effective control of viral transmission^49^. In contrast, point of care tests such as Lateral flow tests (LFT)^50,43^ or those requiring only a basic/mobile laboratory set-up such as Direct RT-LAMP, which have the ability to produce rapid results, can be performed frequently e.g., daily or multiple times per week. Consequently, the likelihood of sampling an individual when their viral load is highest as seen in the early, often pre-symptomatic stages of infection increases, maximising the probability of rapidly detecting infectious cases, allowing prompt isolation. In this use case sampling and testing frequency using a rapid assay with suitable accuracy in detection of medium-high viral loads, but not necessarily optimal sensitivity over the whole range including low to very low viral loads, is desirable or necessary^49,51^. Frequent on-site testing of asymptomatic NHS healthcare workers using Direct RT-LAMP has been successfully implemented in the pilot study described here; and continues to be utilised. Direct-RT-LAMP has also been used in a mass community based pilot in school and higher education settings^36^, to identify those individuals who may have been missed when surveillance relies only on symptomatic individuals coming forward for testing. With the use of mobile or pop-up laboratories, Direct RT-LAMP could also be used for risk-based mass testing, for example, targeting specific geographical areas or vulnerable groups. The potential also exists for lyophilisation of the Direct RT-LAMP reagents reported in other studies^52,53^, which would minimise the necessity for trained personnel by reducing pipetting steps and the requirement for a cold chain, allowing greater capacity of the assay in multi-use case scenarios including point-of-care and in low-and middle-income countries (LMICs).

Several experiments typical of a diagnostic performance evaluation were not performed as part of this study, as they had been performed and reported previously. This included both analytical specificity, which when tested against a panel of respiratory pathogens causing indistinguishable clinical signs to COVID-19, demonstrated a high level of analytical specificity (100% in this case)^3^ and analytical sensitivity of the Direct RT-LAMP, which is reported to detect 1000 cp/ml^3,36,41^. Additionally, the RNA and Direct RT-LAMP assays evaluated as part of this study have been shown to reliably detect the emerging variants of concern (VOC) including the B.1.1.7 alpha variant, the 501Y.V2 beta variant, the P1 gamma variant and the new rapidly spreading B.1.617.2 delta variant^54,55^ (https://www.gov.uk/government/collections/new-sars-cov-2-variant (accessed June, 2021). The emergence of further VOC could lead to a criticism of the RT-LAMP assay due its reliance on a single target, *ORF1ab*, where mutations in the target region in a sample could lead to false negatives. For RT-qPCR this has been observed during the current pandemic ^56– 58^ where at least a dual target assay is recommended^59^. However, this is less likely to occur for the RT-LAMP assay used in this pilot evaluation. Firstly, due to the multiple sets of primer pairs utilised, three pairs, with two pairs within the target region. This builds in redundancy to mutation not unlike a duplex RT-qPCR. Secondly, the ORF1ab region is highly conserved and crucial for viral replication and fitness in SARS-CoV-2. As a result, these regions are well maintained using a proofreading system via the nsp14 protein^60^ resulting in a more stable genome compared to many other RNA viruses.

The authors highlight the importance of incorporating an inhibition control into the next iteration of the RT-LAMP assays. Although the paired RT-LAMP and RT-qPCR data from this study show a good correlation and any false negative results were likely due to the analytical sensitivity of the RT-LAMP assay, not sample driven inhibition. To this end, a control primer set by OptiGene Ltd was evaluated (PS-0010), targeting the human ribosomal protein LO gene. Preliminary analysis of the inhibition control primers showed consistent detection across 279 saliva and 381 combined naso/oropharyngeal swab samples using both RNA and Direct RT-LAMP (manuscript in preparation). Incorporation of this inhibition control into the RT-LAMP assays would alleviate a potential limitation of the current assays and further support quality assurance for use within a clinical diagnostic setting. One further limitation to LAMP assays is the potential for contamination from assay product which can be significant. LAMP assays produce vast amounts which can persist in the environment not only causing potential false positive results in subsequent testing but also anomalous results in laboratory workers who are part of a SARS-CoV-2 testing programme^61^. Therefore, it is crucial that appropriate waste streams are in place to mitigate this risk.

This study demonstrated high sensitivity and specificity for a novel sample preparation method used for SARS-CoV-2 Direct RT-LAMP, particularly in samples from which the individual would likely be considered infectious, highlighting the usefulness of saliva as a simple to collect, non-invasive sample type. The highly sensitive RNA RT-LAMP assay provides a rapid alternative with a reliance on differing reagents and equipment to RT-qPCR testing, thus providing additional diagnostic capacity and redundancy through diversity. Direct RT-LAMP may complement existing surveillance tools for SARS-CoV-2 testing including other point-of-care and laboratory-based diagnostics and is applicable to a variety of clinical scenarios, such as frequent, interval-based testing of asymptomatic individuals that may be missed when reliance is on symptomatic testing alone. However, care should be taken when considering frequency of testing, messaging around the role and interpretation of asymptomatic rapid tests, integration of data storage and access, and the challenges faced when scaling up surveillance to large populations.

The role out of a new testing strategy can often throw up interesting and unexpected experiences. These collective experiences and lessons learnt from setting up an NHS asymptomatic staff testing programme using Direct RT-LAMP will be shared in a future publication.

## Conclusions

Rapid diagnostic testing at scale to identify and isolate symptomatic and asymptomatic individuals potentially transmitting infectious SARS-CoV-2 is an essential part of the response to the COVID-19 pandemic. RT-LAMP on both extracted RNA and directly on crude samples potentially provides faster turnaround times than reverse-transcriptase quantitative real-time PCR testing, with a higher sensitivity and specificity than antigen lateral flow devices. Increasing evidence points to potential benefits of SARS-CoV-2 testing using saliva rather than nasopharyngeal/oropharyngeal swabs, therefore a multi-site evaluation of an improved simple sample preparation method for Direct SARS-CoV-2 RT-LAMP was undertaken. This study demonstrated that the RNA RT-LAMP assay has high sensitivity and specificity, providing a rapid alternative to RT-qPCR testing with a reliance on differing reagents and equipment. The simple SARS-CoV-2 Direct RT-LAMP preparation method also demonstrated high sensitivity and specificity for detecting SARS-CoV-2 in saliva and naso/oropharyngeal swabs from asymptomatic and symptomatic individuals, notably in saliva samples from which the individual would likely be considered infectious. The findings highlight the usefulness of saliva as a simple to collect, non-invasive sample type, potentially applicable for interval-based testing of asymptomatic individuals.

## Data Availability

I have followed all appropriate research reporting guidelines and uploaded the relevant EQUATOR Network research reporting checklist(s) and other pertinent material as supplementary files, if applicable.

## Supplemental Information

**Supplemental Table S1:** Serial dilution of Patient VTM (C_T_ = 19) 1:1 VTM into Lysis Buffer and 98°C heat treatment without and without heat pre-treatment at 56°C for 10 or 30 minutes.

| P07553 ($C_T = 19$) | 1:2 | 1:4 | 1:8 | 1:16 | 1:36 | 1:64 | 1:128 | 1:256 | 1:512 | 1:1024 | 1:2048 |
| --- | --- | --- | --- | --- | --- | --- | --- | --- | --- | --- | --- |
| VTM 1:1 into Lysis + 98°C | D | D | D | D | D | D | D | D | D | D | D |
|  | D | D | D | D | D | D | D | D | D | D | D |
| 56°C 10 mins pre-treat 1:1 VTM into lysis +98°C | D | D | D | D | D | D | D | D | D | D | ND |
|  | D | D | D | D | D | D | D | D | D | D | ND |
| 56°C 30 mins pre-treat 1:1 VTM into lysis +98°C | D | D | D | D | D | D | D | ND | D | ND | ND |
|  | D | D | D | D | D | D | D | D | ND | ND | ND |

D – RNA Detected, ND – RNA Not Detected, by Direct RT-LAMP.

**Supplemental Table S2.** Serial dilution of Patient VTM (C_T_ 19.00 to 32.08) 1:1 VTM into Lysis Buffer and 98°C heat treatment without and without heat pre-treatment at 56°C for 10 or 30 minutes.

| Patient VTM | P07553 ($C_T$ 19) | P01127 ($C_T$ 23.97) | P07102 ($C_T$ 32.08) | P07392 ($C_T$ 24.55) | P01071 ($C_T$ 20.54) |
| --- | --- | --- | --- | --- | --- |
| VTM into 1:1 Lysis + 98°C | D | D | D | D | D |
|  | D | D | D | D | D |
| 56°C 30 mins pre-treat 1:1 VTM into lysis + 98°C | D | D | ND | D | D |
|  | D | D | ND | D | D |

D – RNA Detected, ND – RNA Not Detected, by Direct RT-LAMP.

**Supplemental Table S3.** Sensitivity of the viral culture assay – 1 PFU/ml

| Number of PFU added to AVL/flask | E Gene RT-PCR assay |  |  | Growth (CPE) |  |
| --- | --- | --- | --- | --- | --- |
|  | C<sub>T</sub> value of dilution (AVL) | Baseline Mean C<sub>T</sub> value from flasks | Final Mean C<sub>T</sub> value from flasks | Flask 1 | Flask 2 |
| 1000 | 19.64 | 25.0 | 12.2 | + | + |
| 100 | 22.65 | 28.4 | 11.8 | + | + |
| 10 | 26.00 | 31.8 | 11.9 | + | + |
| 1 | 28.73 | 35.0 | 12.6 | + | + |
| 0.1 | 32.11 | 37.6 | 37.3 | - | - |
| 0.01 | 34.43 | 39.8 | 37.7 | - | - |

### Individual time course

**Supplemental Table S4.** RT-LAMP results of time course from symptom onset

| Days post suspected exposure | Direct RT-LAMP |  |  | RNA RT-LAMP |  |  | RT-qPCR C<sub>T</sub> |  | Observed Symptoms |
| --- | --- | --- | --- | --- | --- | --- | --- | --- | --- |
|  | Tp 1 | Tp 2 | Result | Tp 1 | Tp 2 | Result | ORF1ab | N |  |
| 5 | 09:18 | 09:32 | POS | 11:09 | 10:50 | POS | 23.31 | 26.47 | Onset: Sore throat. Blocked nose. Headache. Lack of appetite. Fever. |
| 6 | 10:32 | 11:34 | POS | 10:34 | 10:40 | POS | 21.16 | 24.03 | Sore throat. Headache. Restless sleeping. Tired |
| 7 | 10:19 | 13:40 | POS | 11:39 | 09:59 | POS | 24.47 | 27.30 | Headache. Restless sleeping. Tired. Loss of smell and taste. |
| 9 | 09:31 | 09:14 | POS | 08:35 | 09:37 | POS | 28.55 | 31.89 | Tired. Loss of smell and taste. |
| 11 | 13:14 | 11:47 | POS | 16:44 | 16:39 | POS | 26.44 | 29.06 | Tired. Loss of smell and taste. |
| 12 | 09:10 | 10:09 | POS | 12:42 | 12:01 | POS | 26.13 | 29.19 | Tired. Improvement in smell and taste. |
| 13 | NEG | NEG | NEG | 14:06 | 12:56 | POS | 28.16 | 30.62 | Significant improvement in all symptoms |
| 14 | NEG | NEG | NEG | NEG | NEG | NEG | 38.05 | 40.73 | None |
| 16 | NEG | NEG | NEG | NEG | NEG | NEG | 36.11 | NEG | None |
| 17 | NEG | NEG | NEG | NEG | NEG | NEG | NEG | NEG | None |

Time to positivity in minutes [Tp]; Cycle Threshold [C<sub>T</sub>]; Negative [NEG]; positive [POS]

